# Evaluation of a Low-threshold Exercise And Protein supplementation intervention for Women (LEAP-W) experiencing homelessness and addiction: Protocol for a single site intervention study

**DOI:** 10.1101/2024.02.29.24303542

**Authors:** Fiona Kennedy, Deirdre Murray, Clíona Ní Cheallaigh, Roman Romero-Ortuno, Julie Broderick

## Abstract

**Background:** Frailty is a complex multi-dimensional state of increased vulnerability to adverse health outcomes and is usually associated with older age but there is growing evidence of accelerated ageing and frailty in non-geriatric populations, including those experiencing socio-economic deprivation and extreme social exclusion, such as people experiencing homelessness. Addiction, as a coping mechanism for prior trauma, is common among people who are homeless and can have a gendered dimension. Women experiencing homelessness and addiction have unique needs which require a gendered approach. The aim of this study is to explore the effectiveness of an exercise intervention to target the known physical functioning deficits and frailty which this population experiences.

**Methods:** This mixed-methods study will explore physical functioning deficits and frailty in women experiencing homelessness and addiction, using a bespoke test battery and an exercise intervention. Physical function (10m Walk Test, 2 Min Walk Test, Single Leg Stance, Chair Stand Test, hand grip dynamometry), frailty (Clinical Frailty Scale and the SHARE- Frailty Instrument) and nutritional status (Mini-Nutritional Status), pain (Numerical Pain Rating Scale) and quality of life (SF 12-V2) will be evaluated. The 10-week intervention will involve a 3-times weekly exercise programme with protein supplementation. Following this, qualitative interviews, which will be thematically analysed using Braun & Clarke methodology, will be conducted. This study will be conducted in Dublin from February to July 2024.

**Discussion:** Little is known about frailty-focussed interventions in women experiencing homelessness and addiction. This proposed study will help to increase the knowledge base regarding the physical health burden and frailty experienced by this extremely vulnerable population and will deliver a targeted intervention with a gendered dimension to mitigate its affects. The findings of this research will help narrow this research gap and will guide clinicians and policy makers to implement unique gender-based treatment strategies for this population. (299 words)

## Introduction

The world’s population is ageing rapidly (1). Frailty, a sometimes-preventable consequence of a rapidly ageing population, is a complex state of cumulative decline across multiple physiological systems which renders a person vulnerable to adverse health outcomes and greatly challenges healthcare systems (2). Targeted interventions to manage frailty in older adults are recommended and exercise with nutritional supplementation has proven effectiveness in combatting and mitigating frailty (3, 4, 5).

Frailty as a construct is a geriatric concept which is strongly associated with increasing age (6). Notwithstanding that biological age is closely linked to frailty, it is known that chronological age alone will not identify frailty as the health and functional life status of populations are so variable (7). Growing evidence indicates the prevalence of premature ageing and frailty in non-geriatric populations (8, 9). ‘Younger frailty’ has been identified in populations of lower socioeconomic status, who live in areas of greater deprivation and experience an earlier onset of illness and disability (10, 11). In a study investigating people experiencing homelessness (PEH) admitted for inpatient care, 83% of participants had mobility problems and 70% were frail or pre-frail despite a low median age of 45 years (12, 13). In an analysis of UK Biobank participants aged 37–73 years, 42% of individuals living with frailty were within the most socially deprived 20% of the cohort (10). Females are known to experience higher levels of frailty compared to males (14, 15). It is therefore not surprising that poor physical functioning and frailty is experienced by young women who are homeless and use drugs (16, 17).

Health services for PEH frequently focus on crisis-based medical, nursing and addiction services, often with a gender-blind lens (18, 19). High levels of stigma, shame and fear result in women accessing services even less and seeking help at breaking point with complex coinciding physical and mental health needs (20, 21). Homelessness and substance use in women is inter-sectoral and may overlap with trauma, abuse, domestic and sexual violence and exploitation, and the needs of these women are complex and unique (22). Pathways into homelessness are frequently different to that of men and require a different approach (23). Internationally, gender-based services for women who are experiencing homelessness and addiction are lacking and services are mostly delivered in mixed gender settings (18, 20). Women-specific services are recommended to increase safety, stability, wellbeing, and psychosocial functioning to respond meaningfully to the serious health and social care needs of women in this setting (24). Gender specific and gender sensitive research is required to fill this knowledge gap (25).

Our prior research, the LEAP I study demonstrated high retention (83% female v 42% males) with excellent engagement and feedback from its female participants, yet caution was observed around engaging in a mixed exercised programme with men (26). Women have traditionally been underserved in research and policy, yet multiple sources cite women as the fastest growing homeless demographic (23). No study has examined how targeted women-only exercise programmes with nutritional supplementation work in practice in this population.

The overall aim of this study is to explore physical functioning and frailty outcomes of an exercise intervention with protein supplementation in a group of (non-geriatric) women experiencing homelessness and addiction challenges. The objectives are:

1. To evaluate recruitment, retention and adherence rates, and any adverse effects of the intervention.
2. To evaluate pre- and post-intervention levels of physical function, nutritional and frailty status, pain, and general health status.
3. To evaluate programme feedback and gain deeper insights into participants perspectives and experiences of the exercise intervention.

## Materials and Methods

### Study design, setting and time frame

This mixed-methods study will involve a single-arm intervention which will take place in Jane’s Place, Merchants Quay Ireland (MQI), in Dublin city centre, a day-service centre which provides services for women who are homeless and/or in addiction. Following the intervention qualitative research in the form of individual exit interviews will be conducted with 20 participants or up to the point of data saturation. This study has received ethical approval from the Faculty of Health Sciences, Trinity College Dublin (Ethical Approval Reference Number: 211202).

### Study population, eligibility criteria and recruitment strategy

#### Study population

This study will involve a non-geriatric population of women who attend Jane’s Place in Dublin city centre. These women have complex needs, many experiencing homelessness and addiction challenges.

#### Eligibility criteria

Inclusion criteria:

➢ Adult women (18-65 years) accessing services in Jane’s Place who consent to participation.

Exclusion criteria:

➢ Any person not meeting the age eligibility criteria.
➢ Any person identifying as a male.
➢ Any person lacking capacity to give consent.
➢ Any person with major physical/medical or cognitive challenges which would preclude ability to safely complete the assessment or
➢ Any person with insufficient English language ability to give fully informed consent.
➢ Women with a confirmed pregnancy.

#### Recruitment Strategy

All potential participants will be provided with a participant information leaflet (PIL) detailing the purpose of the data collection, potential risks and benefits and data protection rights. Where possible a seven-day gap, between receipt of the PIL and obtaining consent, will be provided to allow people time to consider participation. However, if it is their preference to be seen at an earlier and more convenient time this will be facilitated due to the innocuous nature of the evaluation.

Once the research physiotherapist is satisfied that the participant has read and fully understands the PIL, they will proceed to obtain written informed consent. Obtaining consent will take place at the first face to face interaction with the participant prior to commencement of the assessment. The written consent informs participants that they are permitted to withdraw from the study at any time. They will be provided with their own copy of this consent form and PIL with their own signature and that of the research physiotherapist.

#### Stakeholder engagement

Recommendations from the linked LEAP II trial (16) included health education and awareness workshops preceding exercise interventions to enhance health literacy, target lifestyle behavioural change and improve retention rates. Prior to the LEAP- W programme, a number of exercise workshops will be delivered for staff and service users in Jane’s Place. Feedback from this stakeholder engagement will inform the design of the intervention.

### Evaluation and test battery

Using a bespoke data collection form, the following demographic data will be collected: age, gender, ethnicity, living arrangement and environment, highest level of education, marital status, employment status, history of incarceration, history of addiction, self-reported health, and medical history. Physical function, frailty and other related variables will be evaluated using the test battery outlined in Table 1.

**Table 1.** Test Battery.

| Construct Measured | Test |
| --- | --- |
| Sarcopenia and upper limb strength | Upper/Lower Limb circumference<br>Hand grip dynamometry (27, 28) |
| Lower limb strength and endurance | 30-sec Chair Stand Test (29) |
| Endurance | 2-minute walk test (30) |
| Speed | 10m walk test (31) |
| Balance | Single leg stance (32) |
| Frailty | SHARE-FI (33), Clinical Frailty Scale (6) |
| Pain | Numerical Pain Rating Scale (34) |
| Health status/Quality of Life | Short Form 12 (35) |
| Nutritional Status | Mini-Nutritional Assessment (36, 37) |

#### 1. Strength and muscular mass

Muscle strength will be measured, using a Jamar Digital Hand Dynamometer, while sitting with the elbow flexed at 90°, the forearm mid-prone, the wrist in neutral and the hand unsupported (27, 28). Results will be compared to normative reference values (38). Two measurements will be recorded and also used for the SHARE-Frailty Instrument (FI) assessment (33).

Mid-calf circumference girth will be evaluated as this measure correlates with appendicular muscular mass (39). This will be measured using a flexible tape measure at the level of the largest circumference of the calf. Measurements will be compared to gendered cut-off values (40). Mid-arm muscle circumference reflects both muscle mass and caloric and protein adequacy and may be used to signify wasting or malnutrition (41). This test has been recommended in situations where lower limb swelling is present (42). The maximum upper arm muscular mass will be measured using a flexible tape measure. Results will be compared to global reference values (43).

#### 2. Physical performance and lower extremity physical function

This will be measured using the following physical performance measures:

(i) The 10m Walk Test (10mWT). This test measures gait speed and functional mobility and is recorded in m/s. Gait speed is calculated as total distance/time (31).
(ii) The 2 Minute Walk Test (2MWT). This test of self-paced walking ability and functional capacity assesses a participants’ ability to walk unaided over a 15m distance, as fast as possible, for two minutes. Rest breaks are permitted, and the distance covered is measured (30).
(iii) The Chair Stand Test (CST). This tests lower limb strength and endurance and records the total number of sit to stand repetitions performed in 30 seconds (29).
(iv) The Single Leg Stance Test (SLST). This balance test is performed on each leg. The participant is timed standing unassisted on one leg, with eyes open and hands placed on the hips (32).

#### 3. Pain

Each participant will be asked if they are experiencing pain and will be questioned about its location and duration. Pain severity will be assessed using the Numerical Rating Scale (NRS) (34). The NRS is a unidimensional measure of pain intensity from 0-10, with 0 being zero pain and 10 the worst pain imaginable.

#### 4. Frailty

Frailty will be assessed using the Clinical Frailty Scale (CFS) (6) and the SHARE- Frailty Instrument (FI) (33). The CFS is validated for people over 65 years. It is assessed by the tester. Each point on the scale correlates with a level of frailty and a visual chart aids classification from 1 (very fit) to 9 (terminally ill). Higher scores indicate higher levels of frailty. The SHARE-FI is validated for people over 50 years (33). It consists of four brief questions related to the following variables: exhaustion, loss of appetite, walking difficulties and low physical activity, and grip strength measurement. The five results are entered into a freely available web calculator to generate a frailty score and a frailty category of non-frail, pre-frail and frail is also generated. A minor modification was made to the terminology of the SHARE-FI to reflect the real-world experiences of this population.

#### 5. Nutritional status

will be assessed using the Mini-nutritional assessment (MNA) which assesses the risk of malnutrition (36). The short form of the MNA (MNA-SF) (37) is an efficient screening tool consisting of six questions on food intake, weight loss, mobility, psychological stress, or acute disease, the presence of dementia or depression, and body mass index (BMI). The maximum score for this part is equal to 14. A score of 12 or higher indicates a normal nutritional status thus excluding malnutrition and/or malnutrition risk. A score of 11 of less indicates the requirement to proceed with the complete version of the MNA (MNA-LF) (37). The terminology of two of the questions of the MNA (regarding acuity of illness and psychological stress) were slightly modified for the purposes of evaluation of this population in this setting.

#### 8. Short-Form 12 (SF-12)

The SF-12 V2 is a self-report measure of health used across age, disease, and treatment groups (35). It uses eight domains including physical and social activities, pain, mental health, emotional health, vitality and general health perceptions to measure health. The tester will read and complete the 12-question survey with the participant. Results will be entered into a software program provided by the licensing company QualityMetric and two summary scores, mental health (MCS12) and physical health (PCS12) will be generated.

The assessor will document any issues with terminology of the outcome measures utilised, difficulties with completion or floor or ceiling effects reached, as these instruments were designed with geriatric populations in mind.

##### Intervention

This intervention study will be conducted for 10 weeks. It will involve a low threshold, three-times weekly exercise intervention (two exercise classes with protein supplementation and an outdoor ‘Park Walk’). The intervention will be delivered by two research physiotherapists who will adhere to all safety procedures. Flexibility will be facilitated with the provision of a four-week window for pre-intervention evaluations prior to commencement of the programme. Using a trauma-informed lens (20) flexibly arranged group or one-to one sessions will be delivered based on participant preference. Exercise programming strategies and variables are based on our earlier work (LEAP I and LEAP II trials) (16, 17) and also informed by prior stakeholder involvement from people with lived experience. The exercise classes will be multi-modal, with a primary focus on strength and based on core set of resistance exercises (Table 2). Aerobic, balance and flexibility work will be integrated into the class and the exercises will be individualised based on initial assessment results and presentation of participants. Using a gender- based perspective, the following considerations and adaptions will be built into the class; (i) core-stability exercises to target pelvic floor and/or abdominal muscle weakness, (ii) bone building exercises to target peri-and postmenopausal bone loss and (iii) age- associated muscle mass loss. Music, dance and fun orientated physical activity games, an important feature of aforementioned linked studies, will be incorporated to optimize enjoyment and self-esteem. Borg’s Rate of Perceived Exertion Scale will be used to monitor effort and scale the intensity of the workout (44). To promote post-exercise muscle protein synthesis, a nutritional supplement (200ml pre-prepared ‘protein shake’, Fresubin) which consists of 20g of protein will be offered immediately post exercise. The ‘Park Walk’ will focus on the aerobic component of the intervention, using open green space to maximise physical and mental health outcomes. It will be a flexibly arranged 20–30-minute self-paced group or one-to-one walk. To build sustainability beyond the ten-week programme brief health promoting and physical activity educational interventions will be included in the exercise class setting to empower people to engage in unsupervised exercise following the study.

**Table 2.** Exercise Circuit.

| Core exercise | Initial Intensity | Progression/Adaptations* |
| --- | --- | --- |
| Sit to stand/squats/lunges | 2 sets 10-15 reps | 3 sets of 15 reps<br>use of weights/ball |
| Elbow Bends | 2 sets 10-15 reps | 3 sets of 15 reps<br>use of weights/theraband |
| Step-ups | 2 sets 10-15 reps | 3 sets of 15 reps<br>Vary height of step; use of weights |
| Arm elevations or boxing | 2 sets 10-15 reps | 3 sets of 15 reps<br>use of weights or boxing gloves/pads |
| 'Penguin waddle'-hip abduction | 2 sets 10-15 reps | 3 sets of 15 reps<br>additional upper limb abduction and elevation; movement with 360° turns |
| Scapular retractions | 2 sets 10-15 reps | 3 sets of 15 reps<br>Weights/theraband |
| Aerobic activity | 5 mins | Exercise bike, ladders, hurdles, skipping ropes, end of session fun activity. |
| Core set | 5 mins | Pelvic floor exercises; pelvic tilts, bridging, all 4's set |

Adaptations: exercises individualised and progressed for each participant by research physiotherapist
Progression: 2-3 sets of 10-15 repetitions completed at each session; beyond that, low resistance weights (1.1kg-4.5kg), theraband or increased step height introduced to achieve a strengthening effect.

##### Qualitative data

Following the intervention one-to-one in-person exit interviews will be conducted to enable participants to share their feedback and express their views, in order to enhance the understanding of behaviours, adherence and retention challenges and to provide insights for future programmes in this area (45) (S3 Fig). Interviews will continue until 20 participants are seen or until data saturation is reached. The interviews will be audio-recorded and transcribed verbatim and member checking will be utilised.

##### Sample size

As this is a novel field of research, sample size calculations are challenging. The sample method used will be non-probability, convenience sampling, involving the recruitment of a tailored sample that is in proportion to characteristics of the population of interest. This approach may limit generalisability of results but will minimise selection bias (46). A sample size of 30 or greater has been proposed for feasibility studies (47). Considering this and the aim and objectives of this study a target of 40-50 participants has been selected, to allow for drop-out rate of approximately 30%.

## Data collection and management

### Analytic plan

All data will be pseudonymised at point of entry into excel spreadsheets and then transferred into IBM SPSS V28 for analysis. For quantitative data, nominal or ordinal variables will be reported as frequencies and percentages. Continuous variables will be summarised as mean and standard deviation if normally distributed and median and inter- quartile range if non-normally distributed. Data will be tested for normality using the Kolmogorov–Smirnov/Shapiro Wilk test and will be compared across timepoints using the general linear model procedure (normally distributed data) and the Friedman’s test (non-normally distributed data). Chi-squared t-tests will be used where appropriate and some data may be categorised to investigate relationships between variables. Exploratory regression models will be developed to explore correlates and predictors of frailty and poor physical functioning. A *p*-value of <0.05 will be considered significant. For the qualitative data, Braun and Clarke thematic analysis methodology will be employed to provide an in-depth analysis of the exit interview data (48).

## Discussion

Poor physical functioning and frailty has been demonstrated in younger populations who suffer from health disparities and an extreme form of social exclusion, such as PEH and addiction challenges (12, 13, 49). Women who experience homelessness and addiction are especially vulnerable and have unique and complex needs which require a targeted and gender-sensitive approach. Little is known about frailty-focussed interventions in this population, who are often excluded from research and services (18). However, this population have engaged well in two previous linked studies (LEAP I and II). This study aims to address the research gap by providing a targeted exercise intervention to women experiencing homelessness and addiction challenges, thereby providing a gendered dimension, with gendered sensitivity. Its findings will guide clinicians and policy makers in implementing targeted interventions to assist in improving health outcomes in societies most marginalised and vulnerable groups.

## Study status

Recruitment and data collection will commence in February 2024 and will be completed by July 2024.

## Dissemination plans

Conference presentations and publications in peer-reviewed journals will be one method of dissemination. These will be done following the data analysis.

## Acknowledgments

The authors wish to extend their gratitude to the staff of Jane’s Place, MQI who are helping to recruit participants.

## Data availability

There are no underlying data associated with this protocol. Data from this study will be available in open access form.

## Supporting information

S1 Fig. Exit Interview Schedule

